# Unexpected Death of a Duchenne Muscular Dystrophy Patient in an N-of-1 Trial of rAAV9-delivered CRISPR-transactivator

**DOI:** 10.1101/2023.05.16.23289881

**Authors:** Angela Lek, Brenda Wong, Allison Keeler, Meghan Blackwood, Kaiyue Ma, Shushu Huang, Katelyn Sylvia, Ana Rita Batista, Rebecca Artinian, Danielle Kokoski, Shestruma Parajuli, Juan Putra, Chrystalle Katte Carreon, Hart Lidov, Keryn Woodman, Sander Pajusalu, Janelle M. Spinazzola, Thomas Gallagher, Joan LaRovere, Diane Baulderson, Lauren Black, Keith Sutton, Richard Horgan, Monkol Lek, Terence Flotte

## Abstract

An N-of-1 trial was developed to deliver a dCas9-VP64 transgene designed to upregulate the cortical dystrophin as a custom therapy for a Duchenne muscular dystrophy (DMD) patient. After showing signs of mild cardiac dysfunction and pericardial effusion, the patient acutely decompensated and sustained cardiac arrest six-days after dosing and succumbed two-days later. Post-mortem examination revealed severe acute-respiratory distress syndrome with diffuse alveolar damage. Vector biodistribution data was obtained and revealed minimal expression of transgene in liver. There was no evidence of AAV9 antibodies nor of effector T cell reactivity. These findings demonstrate innate immune signaling with capillary leak as a form of toxicity in an advanced DMD case treated with high-dose rAAV gene therapy.

## Introduction

Duchenne muscular dystrophy (DMD) is a fatal, X-linked myopathy caused by mutations in the dystrophin gene, a large structural gene (2.4 Mb genomic DNA with 79 exons)^1,2^ The large size of the DMD gene predisposes this locus to deletions, duplications and point mutations^3^. Dystrophin plays a critical role in the function of cardiac myocytes and skeletal myofibers as part of the dystrophin-glycoprotein complex that anchors myofilaments to the extracellular matrix and prevents stress-mediated damage to the sarcolemma membrane^4,5^.

Several recombinant adeno-associated virus (rAAV)-based approaches to gene therapy for this disorder have been developed^6–8^. The large size of the dystrophin gene has presented a challenge for rAAV-based gene replacement, given the 5-kb packaging limit of the vector capsid, and the need for cis-acting elements, including the AAV inverted terminal repeats (ITRs), a transcriptional promoter and poly-adenylation signal^9^. This issue has led to several innovative approaches including mini-dystrophin^10^ and micro-dystrophin^11,12^ transgenes which include fewer of the internal rod domain repeats. Another approach to dystrophin correction is to use *in vivo* gene editing technologies, often based on the CRISPR-Cas9 system^13,14^. RNA-sequence guided DNA binding can be used to introduce double-strand breaks, as is accomplished with the original Cas9 nuclease^15^. Alternatively, the property of RNA-guiding DNA binding can direct novel engineered Cas9 fusion proteins in which the nuclease activity has been inactivated (“dead“ Cas9 or dCas9) and transcriptional transactivating domains have been introduced^16–18^. This study involved such an approach with dCas9 directing the binding of a VP64 transcriptional activation domain to upregulate a non-muscle full-length isoform of dystrophin (*Dp427c*).

Another critical hurdle for rAAV-gene therapy for DMD is the high dose of rAAV that is required to transduce the extensive mass of tissue that comprises the cardiac and skeletal musculature. Doses of rAAV used in clinical trials for DMD have ranged from 5×10^13^ to 2×10^14^ vector genomes per kg body weight^19,20^. Within this dose range, a number of distinct toxicity syndromes have been observed including hepatotoxicity (often linked to an effector T cell response to capsid or transgene product), thrombocytopenia and thrombotic microangiopathy (TMA), sometimes associated with renal toxicity in an atypical hemolyic uremic syndrome (aHUS) picture, and cardiac toxicity^21^.

The case presented here, while tragic in outcome, presents an opportunity to carefully characterize the systemic, cardiac, and pulmonary toxicities and vector genome biodistribution more fully in the first week to 10 days after high dose rAAV administration. This case also highlights the increased risk for life threatening severe cardiopulmonary failure in patients with advanced DMD when they experience complications of early-phase innate immune activation caused by high-dose rAAV. The short time interval of post-treatment assessments did not allow sufficient time for significant expression of transgene product in the target organs.

### Diagnosis and preclinical studies

The patient was diagnosed with DMD in the first decade of life and has been on daily steroids (deflazacort at 1.1 mg/kg/day) for over 20 years, with loss of independent ambulation occurring in his second decade of life. He had progressive decline in his upper extremity function with increasing cardiopulmonary dysfunction. A skeletal muscle biopsy revealed patchy dystrophin immunostaining (**Figure 1A**), which was shown to be approximately 3% of control muscle by western blot (**Supplementary Figure S1**). Whole genome sequencing showed a ∼30 kb hemizygous deletion encompassing the promoter and exon 1 of the muscle isoform (*Dp427m*) of dystrophin inherited from the maternal genome (**Supplementary Figure S2**). Notably, the deletion leaves the promoter and exon 1 of the cortical (*Dp427c*) and purkinje (*Dp427p*) isoforms of dystrophin intact. We hypothesized that the cortical isoform may be compensating for the absence of the muscle isoform based on previous reports of exon 1 deletions in *Dp427m* observed in X-linked dilated cardiomyopathy cases that result in no overt skeletal muscle phenotype^22,23^. RNA-sequencing results from the patient’s muscle indeed detected low transcript expression of dystrophin derived from the cortical promoter (**Figure 1B**). This finding motivated the design of an individualized therapeutic approach to further upregulate the full-length *Dp427c* isoform, which differs from *Dp427m* in its promoter and exon 1. Using a CRISPR activation approach, we identified the optimal sgRNA to target upregulation of *Dp427c*, first using *in vitro* models, subsequently demonstrating effectiveness in the hDMD/D2-mdx mouse model harboring a humanized DMD locus (manuscript in preparation). **Figure 1C** shows the design of the gene therapy construct, which includes the muscle-specific promoter CK8e and transcription activator VP64 fused to dSaCas9. The size of the transgene insert between the ITRs is 4558 bp and was successfully packaged into the AAV9 serotype. **Figure 1D** shows the timeline of the investigational new drug (IND) preparation by Cure Rare Disease (CRD) in collaboration with Charles River Laboratories, University of Massachusetts Chan Medical School, and Yale University.

**FIGURE 1:**
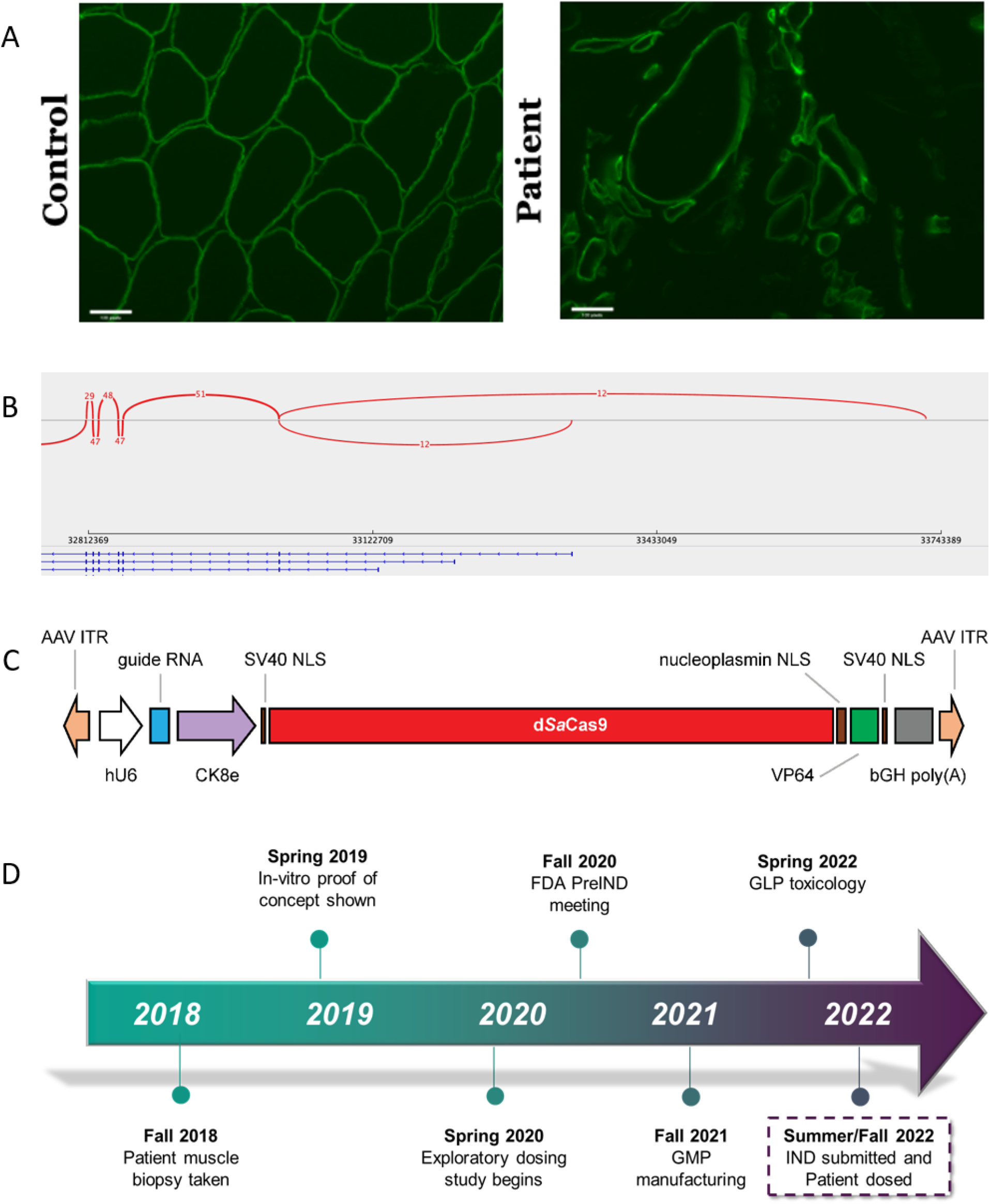
Preclinical assessment and planning towards IND. **A)** Frozen muscle tissue section from a de-identified unaffected individual (left) and the proband (right) immunostained for dystrophin. Dystrophin staining is visible at the myofiber sarcolemma in the unaffected control sample and also in several myofibers of different sizes in the proband. The positive staining was detected using antibodies to the dystrophin rod domain and C-terminus (CAP 6-10 antibody generated in Kunkel Lab). **B)** Bottom panel: RNA sequencing reads showing exons 1–6 (right to left) of the full length *Dp427c* (cortical), *Dp427m* (muscle), and *Dp427p* (Purkinje) DMD isoforms (from top to bottom). Top panel: number of reads that span exons (arcs). Only the DMD cortical isoform is expressed in patient muscle as indicated by reads from *Dp427c* exon 1. **C)** The therapeutic construct was cloned into a plasmid backbone with AAV2 ITRs (Addgene #99680). The guide RNA expression is regulated by a human U6 promoter and the expression of the d*Sa*Cas9-VP64 fusion protein is regulated by a CK8e promoter (Hauscka Lab, University of Washington), which was engineered from the regulatory elements from mouse muscle-type creatine kinase. d*Sa*Cas9, dead *Staphylococcus aureus* Cas9. NLS (nuclear localization sequence), bGH poly(A), (bovine growth hormone polyadenylation signal). **D)** Patient muscle biopsy was performed at University of Massachusetts Chan Medical School in Fall 2018. *In vitro* proof of biology was completed in Spring 2019. Exploratory pharmacology studies performed in collaboration with CRL during Spring 2020. The pre-IND meeting with Food and Drug Administration (FDA) occurred in Fall 2020 and good manufacturing practice (GMP) AAV manufacturing was performed by Andelyn (**Supplementary Table S1**) in Fall 2021 and then good laboratory practice (GLP) toxicology studies commenced in Spring 2022. This culminated in the IND submission in Summer 2022 and patient dosing thereafter.

### Clinical Summary

The patient (in their 20’s) received 1×10-^14^ vg/kg of intravenous CRD-TMH-001 (IND 28497) consisting of AAV9 vector containing CK8e.dSaCas9.VP64.U6.sgRNA. At that time, the patient had severe muscle weakness with a low lean muscle mass of 45%, a restrictive pulmonary defect (FEV1=36% predicted; FVC=36%), and mild left ventricular (LV) systolic dysfunction (LVEF = 54%). He had been on long term steroid treatment (daily deflazacort equivalent to 1 mg/kg/day of prednisone) for over two decades. Baseline immunologic screening showed non-detectable AAV9 total antibody (ELISA <1:25, Athena Diagnostics) and negative ELISPOT responses to AAV9, dSaCas9 (**Figure 2A**). Prophylactic immune suppression therapy was started 13 days prior to dosing (**Figure 2B**). Safety parameters under study included blood counts, serum chemistries, brain natriuretic peptide BNP, and troponin I. Treatment emergent adverse events began 1 day after vector delivery with premature ventricular contractions (PVCs), followed by a downward trend in platelets (2 days post), and increasing BNP (3 days post) (**Figure 2C**). Asymptomatic hypercarbia (pCO2 59 mmHg) with respiratory acidosis was noted on safety monitoring labs 3-4 days post dose. This resolved with optimizing BiPAP pressures from 10/4 to 12/5. Five days post dose the patient developed worsening cardiac function presumed to be myopericarditis given elevation in troponin and pericardial effusion with tamponade physiology. The patient developed sudden acute respiratory distress 6 days post dose, with CXR findings of acute respiratory distress syndrome (ARDS) and worsening cardiac function (EF 45–50%). The patient progressed to cardiopulmonary arrest and was emergently placed on extracorporeal membrane oxygenation (ECMO). Despite support with ECMO, the patient passed away 8 days post-treatment due to multiorgan failure and severe neurological injury. Laboratory studies from the post-vector period indicated high IL6 (2.8 ng/mL) and a mixed picture of complement components with elevated C5b-9. Multiplex cytokine bead-based assays revealed elevations of IL-8 in the serum (**Supplementary Figure S3**) and high levels of IL-6 and MCP-1 in the pericardial fluid (**Supplementary Figure S4**). Mitigating therapies attempted during this time included increased steroids, eculizumab (anti-C5), tocilizumab (anti-IL6-R), and anakinra (IL1-R blocker).

**FIGURE 2:**
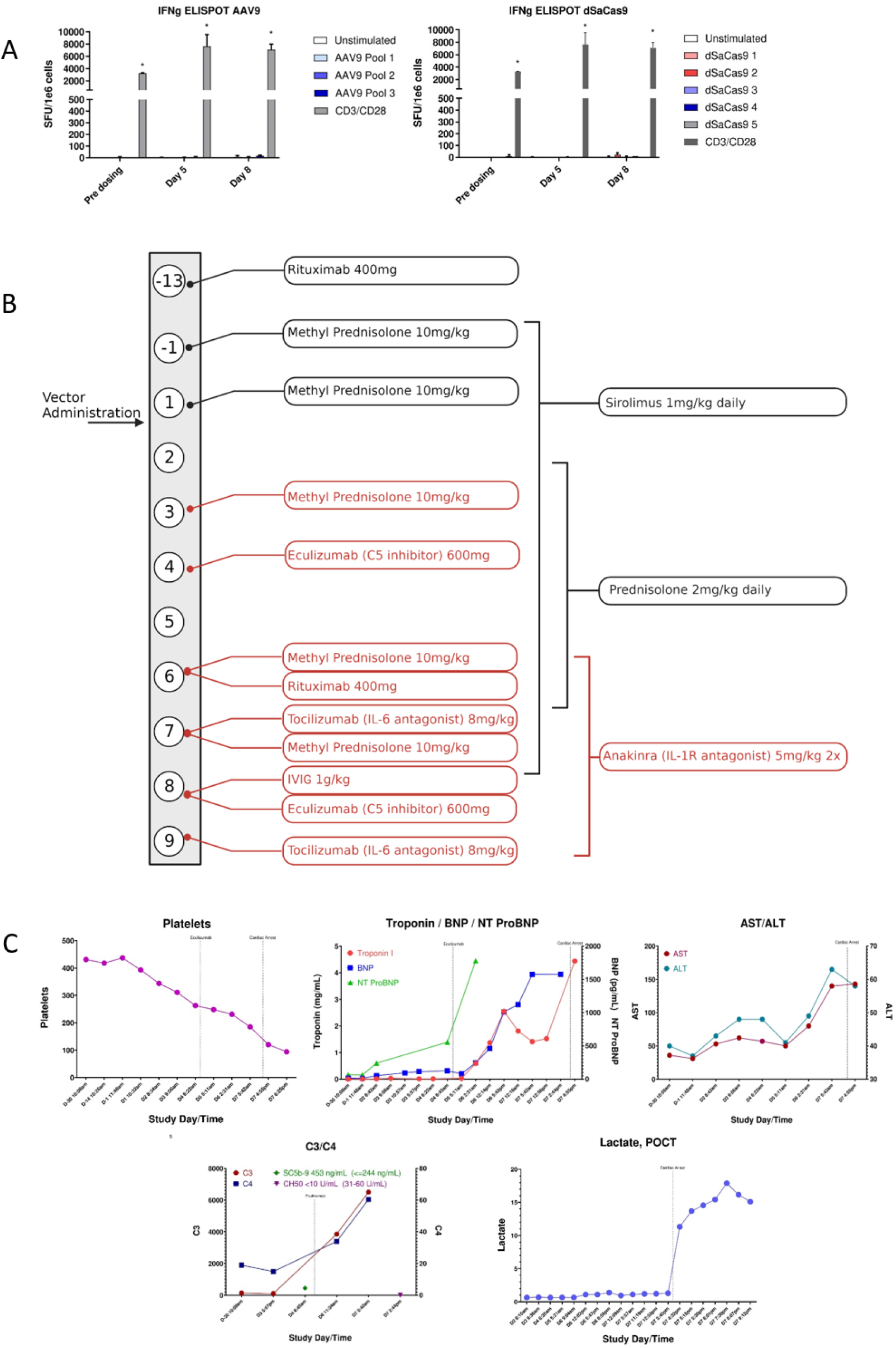
Clinical trial data. **A)** Interferon-gamma ELISPOT assays for AAV9 and SaCas9 in patient samples. PBMCs isolated from the patient at different time-points in life were stimulated by either AAV9 (left) or dSaCas9 (right) peptide pools to assess T-cell responses by interferon-γ ELISPOT assay. Negative controls were unstimulated cells, and CD3/CD28 stimulation was used as positive control. Data was run in technical triplicates and reported as mean ±SD. Significance designated by * represents 3x negative control. Peripheral blood mononuclear cells (PBMCs), spot forming unit (SFU) standard deviation (SD). **B)** Timeline of prophylactic immune suppression administered during clinical trial. Numbered circles denote day of treatment - beginning 13 days pre-treatment to 9 days post-treatment. Drugs shown in black text were listed on the clinical trial protocol, drugs shown in red were not. **C)** Monitoring of cardiac, complement and liver markers during clinical trial period (beginning 30 days prior to vector administration).

### Post-mortem Studies

A consent for limited autopsy allowed for gross and microscopic examinations of the heart, lungs, brain, triceps, and liver. The post-mortem examination confirmed markedly decreased muscle mass in the heart and skeletal muscle. Examination of the heart demonstrated severe cardiomyopathy, characterized by significant gross and histologic fibrofatty replacement of biventricular myocardium (**Figure 3A**), consistent with what has been described in dystrophin-deficient cardiomyopathy^24^, and without overt features of active inflammation/myocarditis, thrombotic microangiopathy, or complement deposition (confirmed by immunohistochemistry, **Supplementary Figure S5)**. The lungs were heavy and edematous (600 gm combined weight, compared with 475 gm expected); the histology showed diffuse alveolar damage, characterized by hyaline membrane formation along with interstitial and intra-alveolar edema (**Figure 3B**). The findings were in keeping with the clinical impression of ARDS. There was no evidence of thrombotic microangiopathy or significant inflammation. Gross and microscopic examinations of the brain demonstrated infarctions in a “watershed” distribution in the cerebral cortex and cerebellum, and widespread neuronal injury likely reflecting poor perfusion in the pre-terminal stage (**Supplementary Figure S6**).

**FIGURE 3:**
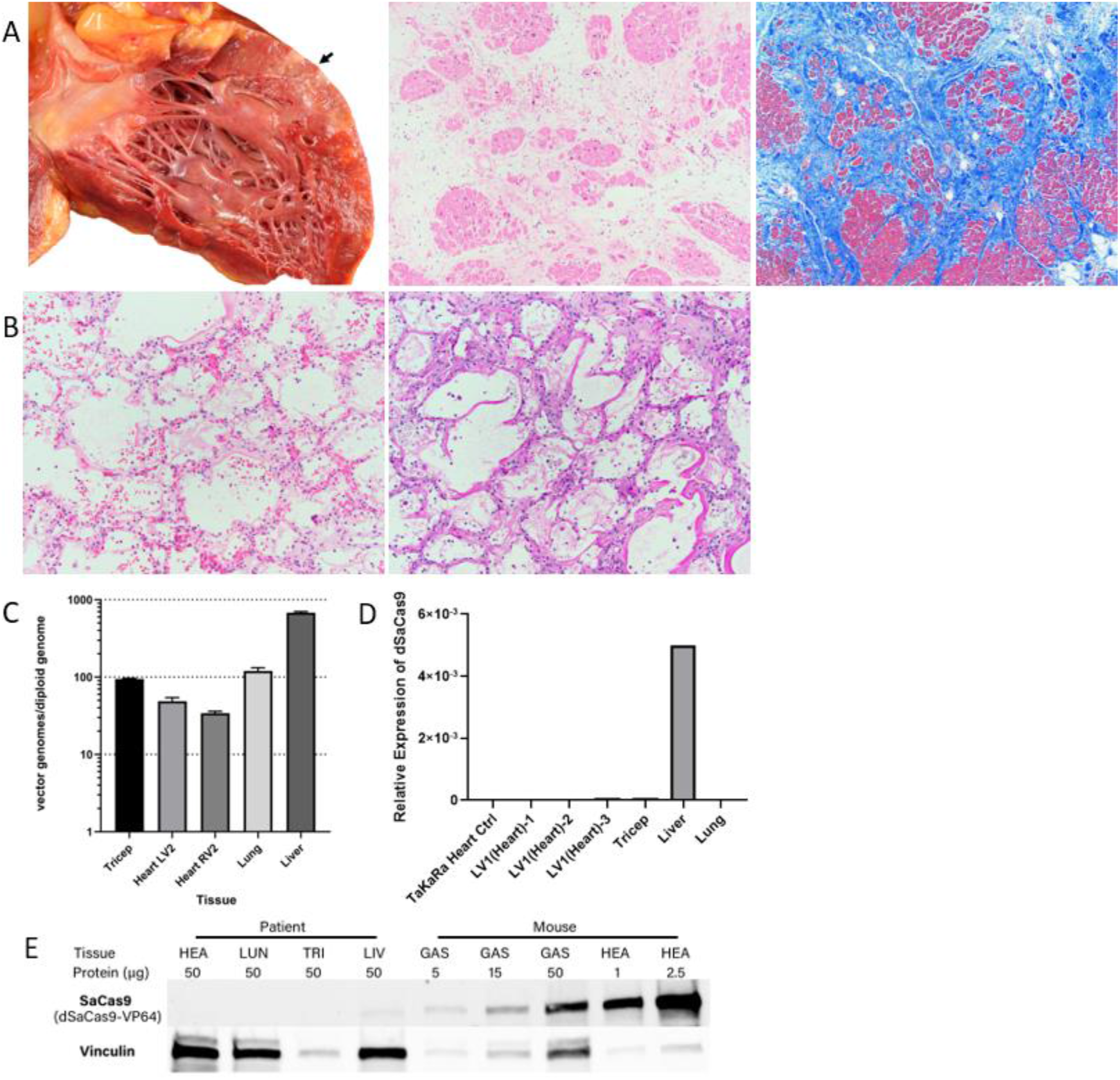
Post-mortem analysis of patient tissues. **A)** Macroscopic examination of the heart demonstrates fibrofatty replacement of the left ventricular wall (arrow); histologic evaluation of this area shows marked interstitial fibrosis and fatty replacement with residual cardiac myocytes; there is no histologic evidence of myocarditis or thrombotic microangiopathy (H&E and Masson trichrome stains, 4X). The findings are in keeping with severe cardiomyopathy. **B)** Microscopic examination of the lungs shows diffuse alveolar damage, characterized by hyaline membrane deposition with interstitial and intra-alveolar edema (H&E and PAS stain, 10X). **C)** Vector biodistribution in patient tissues. Vector genomes were quantified by qPCR and calculated to indicate vector genomes per diploid genome for each tissue. Data was run in technical triplicates and reported as mean ±SD. Quantitative polymerase chain reaction (qPCR) standard deviation (SD). **D)** RNA expression levels of d*Sa*Cas9 were measured by RT-qPCR. Heart tissue was sampled in three different locations. **E)** SaCas9 expression in post-mortem tissue compared to AAV-injected mice at similar dosage. Protein expression in GAS and HEA mouse tissue is from 8-week time point compared to 8-day post-treatment from patient tissues. Vinculin was used as a loading control but showed variable relative expression in non-muscle tissues. HEA, heart; LUN, lung; TRI, triceps; LIV, liver; GAS, gastrocnemius.

### Vector Biodistribution

Analyses of AAV vector DNA distribution were performed using quantitative real-time PCR (qPCR) (**Figure 3C**). Vector genomes were detected in lung tissue at a level of 119 vg/diploid genome. Likewise, vector genomes were detected in the myocardium, with 34 vg/diploid genome in the left ventricle sample and 48 vg/diploid genome in the right ventricle sample. Vector genome abundance in liver indicated 680 vg/diploid genome. Digital droplet PCR (ddPCR) was used to confirm vector genome analysis but genome abundance in liver exceeded upper limit for accurate quantification (**Supplementary Figure S7**).

### Expression of Transgene Products

SaCas9 transcript expression was assessed using qPCR and revealed no detectable levels in the tissues analyzed except for liver (**Figure 3D**). This pattern was also observed for SaCas9 protein, as only a faint band was detectable in the liver by western blot (**Figure 3E**). The absence of the transgene in skeletal and cardiac tissues did not warrant measurement of *Dp427c* upregulation at this timepoint post-mortem. Expression levels of guide-RNA showed similar tissue trends (**Supplementary Figure S8**). No AAV9 capsid or Cas9 transgene specific T-cell responses were detected by interferon gamma ELISPOT in patient’s PBMCs at days 4 or 7 post-dosing (**Figure 2B**).

## Discussion and Conclusions

We present the case of an advanced stage DMD patient who experienced severe cardiopulmonary toxicity within 6 days following IV administration of rAAV9-dCas9VP64 at a dose of 1×10^14^ vg/kg. Per our clinic-pathologic findings, we hypothesize this patient developed a cytokine-mediated capillary leak syndrome, manifested by pericardial effusion on day 5 and rapidly developing ARDS on day 6. The latter resulted in acute worsening of pulmonary compliance with respiratory failure, hypoxemia, and an associated acute worsening of right ventricle heart failure. Unlike other DMD patients in rAAV trials, this patient did not exhibit evidence of TMA or of adaptive humoral or cell-mediated immune responses to AAV capsid or transgene products.

Unfortunately, the acute toxicity and shortened course of this patient prevented a substantive assessment of the safety and efficacy of the CRISPR-transactivator approach itself. It is well-known that the single-stranded rAAV genome requires weeks to form transcriptionally active double-stranded forms after *in vivo* gene therapy. In this case, the innate immune toxicity shortened the patient’s course to an extent that would not have been predicted to allow for significant transgene expression. While trace amounts of the transgene product were present in the liver, none was detectable from cardiac or skeletal muscle, and no effector T cell responses to dCas9 or AAV9 were observed. When comparing vector biodistribution results in our patient against two patients dosed with systemic administration of Onasemnogene abeparvovec (AAV9) for Spinal Muscular Atrophy^25^, the vg per diploid genome in the liver are comparable; but notably higher in the heart, lung, and muscles in our patient; which may be due to the short duration post-injection. It is plausible that the extensive loss of myofibers in the patient may have altered the expected vector biodistribution to these tissues. However, this higher vector genome load in our patient occurred despite dosing at 1×10^14^ vg/kg dose (relative to 2×10^14^ vg/kg for microdystrophin DMD clinical trials) due to his lower lean muscle mass of 45%. Dose determination will remain a challenge for custom-designed AAV-mediated therapies, as by definition the precise therapeutic dose will not have been established.

Another recent fatal case was described in a trial involving a non-ambulatory 16-year-old DMD patient, who passed six days after receiving gene therapy^21^. The patient was part of Pfizer’s gene therapy trial (NCT03362502) and received fordadistrogene movaparvovec at 2×10^14^ vg/kg. His death is believed to be linked to an innate immune response against the capsid in the myocardium, which led to cardiogenic shock and heart failure. In our case, we suspected myocarditis clinically because of an acute rise in serum troponins and pericardial effusion. However, the opportunity for direct histopathological examination of the myocardial tissue post-mortem gave definitive evidence of the absence of innate immune cell infiltration typically seen in myocarditis.

There are several novel aspects of our case that add to the body of knowledge relevant to future applications of AAV-mediated gene therapy to DMD; however, our interpretations and separation of what is due to the custom gene therapy administered, age of the patient, and severity of disease state is challenging because of the design of this trial with a unique AAV-mediated gene therapy for a single patient. The patient’s late stage of DMD progression at dosing may have limited his physiologic reserves to tolerate the cardiopulmonary stress associated with acute toxicity resulting from rAAV gene therapy, but this was not possible to fully demonstrate. Capillary leak syndromes have been well-described following cytokine release in other gene and cell therapies, including systemic rAd and CAR-T cell therapies, but not typically after higher doses of AAV for rare genetic diseases. To our knowledge, this is the first reported case of severe ARDS in AAV gene therapy in DMD. It is possible that this is unique to the specific product; however, since the toxicity ensued prior to detectable levels of transgene expression and since the protein composition of the vector preparations was equivalent to other high-dose AAV9 vectors, it is more likely that this was host-specific rather than vector-specific. Our interpretation is that the patient described here appears to have experienced a more severe innate immune reaction than others receiving similar or slightly higher doses of rAAV in microdystrophin gene therapy trials; thus, further research into host characteristics predisposing to severe innate immune reactions to AAV may broadly improve safety of AAV-mediated gene therapy at high doses. As more applications of high-dose IV rAAV gene therapy are developed the potential for such toxicities should be considered and carefully monitored among patients whose underlying disease may lessen their ability to tolerate these adverse effects, especially for custom-designed gene therapy products without prior dosing in humans.

## METHODS

Preclinical assessment was performed on patient quadricep muscle biopsy and dystrophin protein quantification was performed by western blot and immunostaining. RNA was extracted to perform RNA-sequencing and assess expression of *DMD* transcripts. Whole genome sequencing was performed on the patient and mother using blood-derived DNA.

The patient provided written informed consent for the clinical trial study which was sponsored by Cure Rare Disease and performed in accordance with protocols approved by the institutional review board at University of Massachusetts Chan Medical School. Clinical-grade plasmid and vector were manufactured at Aldevron and Andelyn, respectively. The trial was conducted at University of Massachusetts Chan Medical School; imaging and safety labs were performed at U Mass Memorial Hospital Medical center; AAV9 antibody testing was done at Athena Diagnostics.

A limited autopsy was consented by the patient’s family in which liver, brain, skeletal muscle, lungs, heart tissue were evaluated by the pathology department of Boston Children’s Hospital. Postmortem examination was performed 19 hours following death. Macroscopic and microscopic examination was conducted for each organ. Routine H&E stains were evaluated for representative sections of each organ. Special stain (Masson trichrome for fibrosis) and immunohistochemical stains (CD3, CD20 and C4d for T cells, B cells, and complement deposition, respectively) were performed in selected sections of cardiac tissue. Periodic acid-Schiff (PAS) stain was performed on selected lung sections to confirm hyaline membrane deposition. In addition, DNA, RNA and protein were extracted from tissues for vector genome copy number, transgene transcript and protein quantification, respectively **(Supplementary Method)**.

## Data Availability

All data produced in the present study are available upon reasonable request to the authors

## Notes

### Competing Interest Statement

The authors have declared no competing interest.

### Clinical Trial

IND28497
NCT05514249

### Funding Statement

This study was funded by the 501c3 foundation Cure Rare Disease.

### Author Declarations

The protocol was reviewed and approved by the UMass Chan Medical School Institutional Review Board (IRB), Institutional Biosafety Committee, and Center for IRB Intelligence (CIRBI). External safety oversight was provided by an independent data safety monitor. The study was approved by the Food and Drug Administration (IND 28497). Families gave written informed consent for the study.

### Summary of Updates

There was an error in the units reported for IL6. It has now been corrected to 2.8ng/ml.

