## Supplementary Appendix for "Unexpected Death of a Duchenne Muscular Dystrophy Patient in an N-of-1 Trial of rAAV9-delivered CRISPR-transactivator"

### TITLE

### SUPPLEMENTARY METHODS

#### Western blot on human muscle biopsies

Snap-frozen normal and DMD patient quadricep muscle biopsies were ground using a mortar and pestle over dry ice and lysed in RIPA buffer (50 mm HEPES pH 7.5, 150 mm NaCl, 5 mm EDTA, 1 mm EGTA, 15 mm p-nitrophenyl phosphate disodium hexahydral, 1% NP-40, 0.1% SDS, 1% deoxycholate and 0.025% sodium azide) with protease and phosphatase inhibitor cocktails added (Sigma). Lysates were incubated on ice for 1 h, vortexing halfway through, centrifuged at 16000g for 30 min at 4°C and the supernatants retained. Protein concentration was measured using a BCA protein assay kit (Bio-Rad). Lysates were separated by SDS–PAGE on 4-20% Tris–glycine gels (Bio-Rad) and transferred to PVDF membranes (Life Technologies). Membrane was blocked in 5% milk and 2% BSA in TTBS and then probed with rabbit polyclonal Dystrophin 6-10 antibody (Lidov, *et al*., 1990) at 1:750 followed by HRP-linked rabbit secondary (Cell Signaling, 7074). Blot was developed with Pierce ECL Plus (Thermo Scientific, 32132) on Blue Devil autoradiography film (Genesse Scientific, 30-101). Western blot was analyzed by ImageJ software.

#### Transcript expression analysis on post-mortem tissues

**Primers:**

hHPRT1-F: CATTATGCTGAGGATTTGGAAAGG

hHPRT1-R: CTTGAGCACACAGAGGGCTACA

129 bp

SaCas9_F: CTACGAGGCCAGAGTGAAGG

SaCas9_R: TCGGCCACGTATTTCTCTTC

192 bp

C7-F: GATCATGCGAAAGGGGAGC

C7-R: TCTCGCCAACAAGTTGACG

98 bp

**Standard curves:**

Standard Value (dSaCas9) = 4006.5765672931*e^(-0.6374394874*Ct)

Standard Value (hHPRT1) = 6720571.7317989400*e^(-0.6080430546*Ct)

C7 standard curve was not generated due to low expression.

**Definition of expression levels of standard curves:**

The dSaCas9 expression in 200 ng RNA from the heart tissue of mouse 3011 (clinical construct injected hDMD/D2-mdx mouse) and the hHPRT1 expression in 200 ng TaKaRa Human Heart RNA was defined as 1 for their standard curves, respectively.

**RNA isolation**

Tissues were added to 1 mL of TRIzol reagent (Life Technologies, 15596018) and homogenized until completely dissolved using a tissue homogenizer (Omni International, 10046-846). Once homogenized they were incubated at room temperature (RT) for 5 minutes (mins) to permit further dissociation. 200 µL of 1-Bromo-3-chloropropane (VWR, 10841- 634) was added and samples were vortexed vigorously then incubated at RT for 2-3 mins. Samples were centrifuged at 12,000 x g 4 °C for 15 mins to separate the RNA, DNA and protein phases. The aqueous RNA phase was carefully removed into a new 1.5 mL microcentrifuge tube and 500 µL of isopropanol (Sigma Aldrich, I9516) was added to precipitate the RNA. Samples were inverted to mix and incubated at RT for 10 mins following subsequent centrifugation at 12,000 × g for 15 mins at 4°C. The supernatant was removed from the pellet and the pellet was washed twice in 75% ethanol and centrifuged at 10,000 × g for 10 mins at 4°C. The supernatant was removed and the pellet was air dried for 10 mins at room temperature. The RNA pellet was resuspended in nuclease free water depending on pellet size (50–800 µL). Concentration and purity were measured using NanoDrop (Thermofisher Scientific, ND2000).

**DNase Treatment**

Residual genomic DNA in the RNA samples were removed using the Invitrogen ezDNase kit (Thermofisher Scientific, 11766051). Briefly, 2 µg RNA samples were diluted to 8 µL with Nuclease free water. 1 µL ezDNase and 1 µL reaction buffer were used for each reaction. The samples were incubated at 37°C for 10 mins and then the DNase was deactivated at 70°C for 3 mins in a ProFlex thermal cycler (Applied Biosystems, 4484073).

**One-step RT-qPCR**

The RT-qPCR reaction used 20 ng RNA (dSaCas9) or 200 ng RNA (hHPRT1; C7) as a template and the reaction contained iTaq Universal SYBR Green Reaction mix and Reverse transcriptase (BioRad, 1725151), forward and reverse primers (final concentration of 250nM each) and nuclease free water to a total volume of 20 µl. The qPCR reaction was prepared in Multiplate 96 well PCR plates (low profile) (BioRad, MLL9601) which were sealed using Microseal 'B' PCR Plate Sealing Film, adhesive (BioRad, MSB1001).

| **Reagent** | **Amount** |
|---|---|
| 20 ng RNA or 200 ng RNA | x µL |
| 10 µM Forward & Reverse primer mix | 1 µL |
| Reverse transcriptase | 0.25 µL |
| Reaction mix | 10 µL |
| nuclease free water | Up to 20 µL |

The qPCR was performed on a CFX96 C1000 touch real time PCR detection system (BioRad, 1855196) using a 2-step amplification plus melt curve with the following parameters. qPCR triplicates were performed for each reaction.

| Temperature | Time | Number of Cycles |
|---|---|---|
| 50 °C | 10 mins | x1 |
| 95 °C | 3 mins | x1 |
| 95 °C | 10 secs | x39 plus plate read |
| 57 °C | 1 min | |

To check for primer specificity an additional melt curve analysis step was performed starting at 65°C for 0.05 seconds with 5-degree increments until 95 °C with a plate read step to capture data.

For data analysis Ct values were collected. The expression was quantitated using the standard curves (dSaCas9) or ddCT (C7).

**Protein Extraction and Quantitation**

Mouse tissues were harvested 4 weeks post-injection. Both human and mouse tissues were homogenized in 250–500 µL of ice-cold protein extraction lysis buffer made by adding 10 µL of protease inhibitor cocktail (Sigma Aldrich, P8340-1ML) to 1 mL of RIPA lysis and extraction buffer (Pierce, PI89900). After 20 minutes of incubation on ice, samples were vortexed again, and sonicated by Bioruptor® Plus Sonication System (Diagenode, B01020001) for 13 rounds of 30s on and 30s off. Samples were centrifuged for 30 minutes at 16,000 × g at 4 °C in a refrigerated centrifuge (Eppendorf, 5404). The supernatant was transferred to a clean 1.5 mL microcentrifuge tube and placed on ice until progressed to the quantitation.

Protein was quantitated using the BioRad DC Assay II kit (BioRad, 5000112). Using the BSA reagent from the kit, seven standards are prepared from 0–1.5 mg/mL in the protein extraction buffer from above for creating a standard curve. Measurements were done according to the instruction manual, and absorbances at 750nm were read on a plate reader (BioRad, #1681135). Concentration is calculated relative to the standard curve.

**SDS-PAGE and western blotting for mouse and post-mortem tissue**

Appropriate amount of protein extraction lysis buffer from above was added into a specific loading amount of each protein sample to achieve an equal volume for each sample, which was then prepared in 4x Laemlli sample buffer (BioRad, 1610747). Samples were mixed by a brief vortex and then heated at 95°C on a heat block (BioRad, 1660571) for 5 minutes. Samples were cooled down on ice and then centrifuged briefly at max speed to ensure equal loading (Eppendorf, 5404). 3.5 µL of PageRuler Plus prestained protein ladder (ThermoFisher Scientific, 26619) and all of the samples were then loaded onto a 4-15% mini-protean TGX stain free protein gel (BioRad, 4568084) and placed into the Mini-PROTEAN tetra cell electrophoresis system (BioRad, 1658000). SDS-PAGE was performed at 100V for 80 minutes using a PowerPac HC power supply (BioRad, 1645052). Upon completion of the SDS-PAGE, the gel was removed and transferred onto nitrocellulose membrane (BioRad, 1704158) by the TransBlot Turbo System (BioRad, 1704150EDU) using the 7-minute transfer program (2.5 A constant, up to 25 V variable).

Membrane blocking buffer was made of 5% Omniblock non-fat dry milk (American Bio, AB10109-00100) in 1× TBS-Tween 20 (TBS-T). The membrane was then blocked for 1 hour in membrane blocking buffer. Primary antibody Anti-CRISPR-Cas9 (Abcam, ab203943) was made up 1:1,000 in membrane blocking buffer and the membrane was incubated in the primary antibody overnight at 4°C whilst rocking to equally distribute antibody (BenchRocker 2D Genesee Scientific, 31-201). The following day the membrane was washed 5 times for 5 minutes in TBS-T and incubated in secondary anti-rabbit HRP conjugated secondary antibody (Cell Signaling Technology, 7074S) at 1:2000 in membrane blocking buffer for 2 hours at room temperature. Membrane was washed again 5 times for 5 mins at room temperature in TBS-T prior to developing with Clarity Max ECL substrate (BioRad, 1705062) using the Chemidoc Imaging system (BioRad, 12005606). Both chemiluminescent and colorimetric images were taken to observe protein expression and size of bands. To determine equal protein loading, the membrane was stripped for 15 minutes in Restore PLUS western blot stripping buffer (ThermoFisher Scientific, 46430) at room temperature, washed in 1x TBS-T and then re-blocked in membrane blocking buffer for 1 hour. Vinculin was used as a loading control, thus the membrane was incubated in primary antibody (Sigma Aldrich, V9131-100 µl) at 1:80,000 dilution overnight at 4 °C whilst rocking to equally distribute antibody. Washes and secondary were performed as described using Anti-mouse HRP conjugated secondary antibody (Cell Signaling Technology, 7076S). Membrane was developed as previously using Clarity ECL substrate (BioRad, 1705060) and the Chemidoc Imaging system (BioRad, 12005606).

**CBA assay**

For the detection of cytokines in serum, Cytometric Bead Array (CBA) assay was performed using Biolegend LEGENDplex HU Essential Immune Response Panel (13-plex) with V-bottom Plate (Biolegend, Cat# 740930) or Biolegend LEGENDplex Human Type 1/2/3 Interferon Panel (5-plex) with V-bottom Plate (Biolegend, Cat# 740396) following the manufacturer's recommendations. For the detection of cytokines in peri-cardiac fluid, Cytometric Bead Array (CBA) assay was performed using Biolegend LEGENDplex Human Type 1/2/3 Interferon Panel (5-plex) with V-bottom Plate (Biolegend, Cat# 740396) following the manufacturer's recommendations. Serum samples from the patient were diluted 1:2. The peri-cardiac fluid sample from the patient was not diluted for the assay. Data were acquired on a BD LSRII and analyzed using the LEGENDplex Data Analysis Software Suite.

**ddPCR**

Digital droplet PCR (ddPCR) was performed using 5ng of gDNA and a human RPP30 reference primer/probe as well as the following primers and FAM labeled probe for the CK8e promoter:

Forward primer: accctgcatgccatgttc

Reverse primer: ctgacttgctcactggttcc

Probe: cccgccagctagactcagca

Samples were run on the Bio Rad QX200 system according to the manufacturers recommendations. To be considered valid, each well is required to have at least 10,000 accepted droplets, of which 100 must be negative.

**qPCR for vector biodistribution**

To measure vector biodistribution, harvested tissue was processed for genomic DNA, using the Gentra Puregene Kit (Qiagen) as per manufacturer's recommendations. Vector genome copy numbers were measured by qPCR in 20 ng of total DNA, using the following primers and probes for the BGH polyA in the vector:

forward primer CCTCGACTGTGCCTTCTAG;
reverse primer TGCGATGCAATTTCCTCAT;
probe 56-FAM/TGCCAGCCA/ZEN/TCTGTTGTTTGCC/3IABkFQ.

All samples were compared to a plasmid-based standard curve and a viral vector internal control for quality assurance. The primer-probe mix was acquired from IDT, and the reaction was run using TaqMan Gene Expression Master Mix (Applied Biosystems) in a CFX Opus 96 qPCR system (Bio-Rad).

**ELISPOT**

An ELISpot assay detecting the secreted INFg was performed on patient PBMCs to measure the cellular immune response to the AAV9 capsid and dSaCas9 transgene. Peptides spanning the transgene and the VP1 of the capsid, consisting of 15-mers that overlap by 10aa (mimotopes), were divided separately into pools (five pools for the transgene and three pools for the capsid). The cells are stimulated for 48hrs with the peptides, a negative control (media only), and a positive control (CD3/CD28, Mabtech) for cytokine secretion. Spots numbers were analyzed and counted using the Mabtech IRIS reader. To consider the cellular immune response positive, the number of spot-forming units per $10^6$ cells must be greater than 50 and 3-fold higher than the CD3/CD28 positive control.

### SUPPLEMENTARY FIGURES

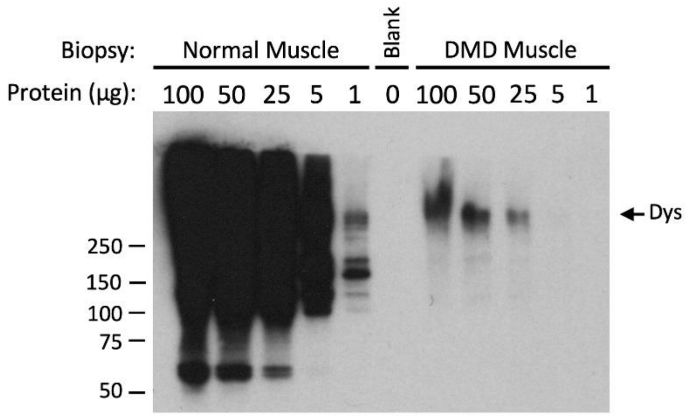

| Lane | Normal (PD) | DMD (PD) | Normal Adjusted (PD) | (DMD/Normal Adjusted)*100 | Average % | Stdev | SE |
|---|---|---|---|---|---|---|---|
| **1µg Normal** | 7971.823 | | | | | | |
| **100µg DMD** | | 20485.409 | 797182.300 | 2.570 | | | |
| **50µg DMD** | | 12675.581 | 398591.150 | 3.180 | **2.879** | 0.305 | 0.176 |
| **25µg DMD** | | 5752.225 | 199295.575 | 2.886 | | | |

**Figure S1: Dystrophin expression in normal and DMD patient muscle.** Top: Western blot of normal and DMD patient quadricep muscle biopsies shows significantly diminished dystrophin expression in DMD patient compared to normal muscle. Bottom: Western blot quantification indicates ~2.9% dystrophin protein expression in DMD patient compared to normal muscle. PD = pixel density as quantified in ImageJ.

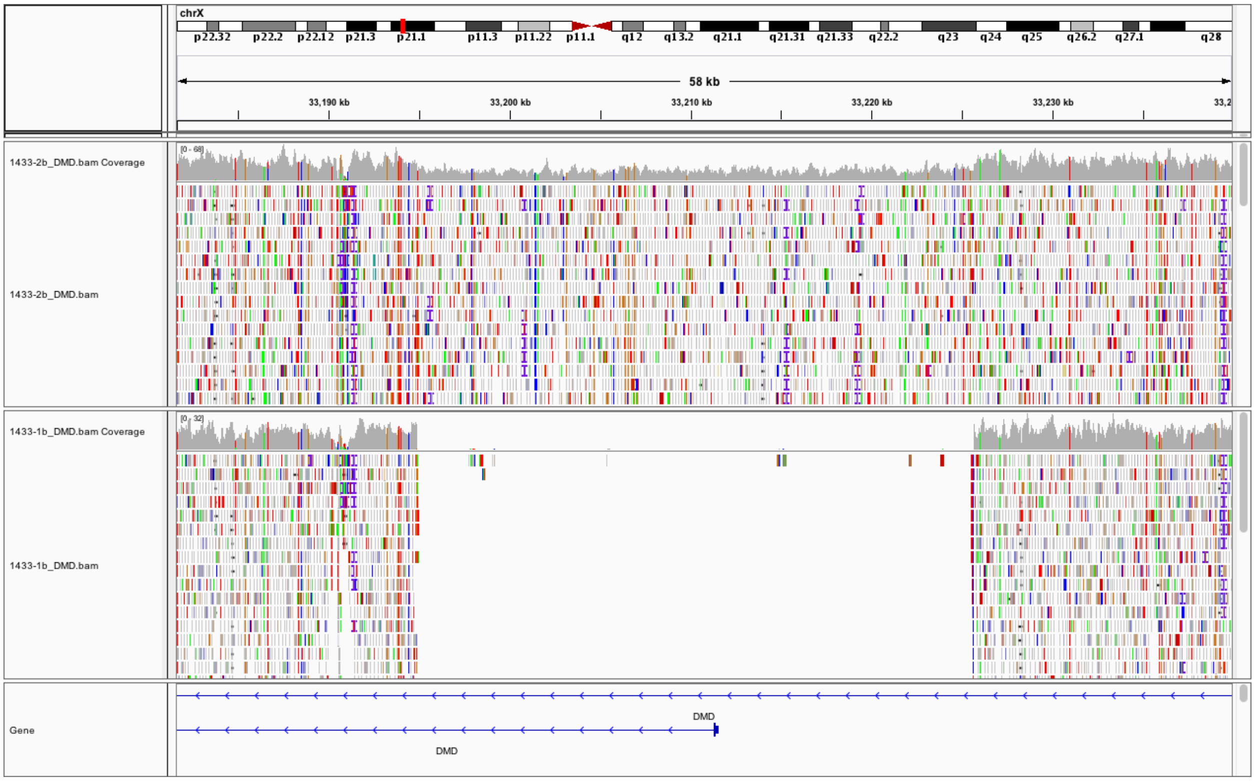

**Figure S2: Whole genome sequencing of patient and mother DNA.** Whole genome sequencing reveals a 30 kb hemizygous deletion (GRCh38 X:x-y) encompassing the promoter and exon 1 of the muscle (*Dp427m*) isoform (bottom panel). The mother of the patient (top panel) is heterozygous for the deletion revealed by 0.5× coverage in the histogram in the overlapping region and split read support.

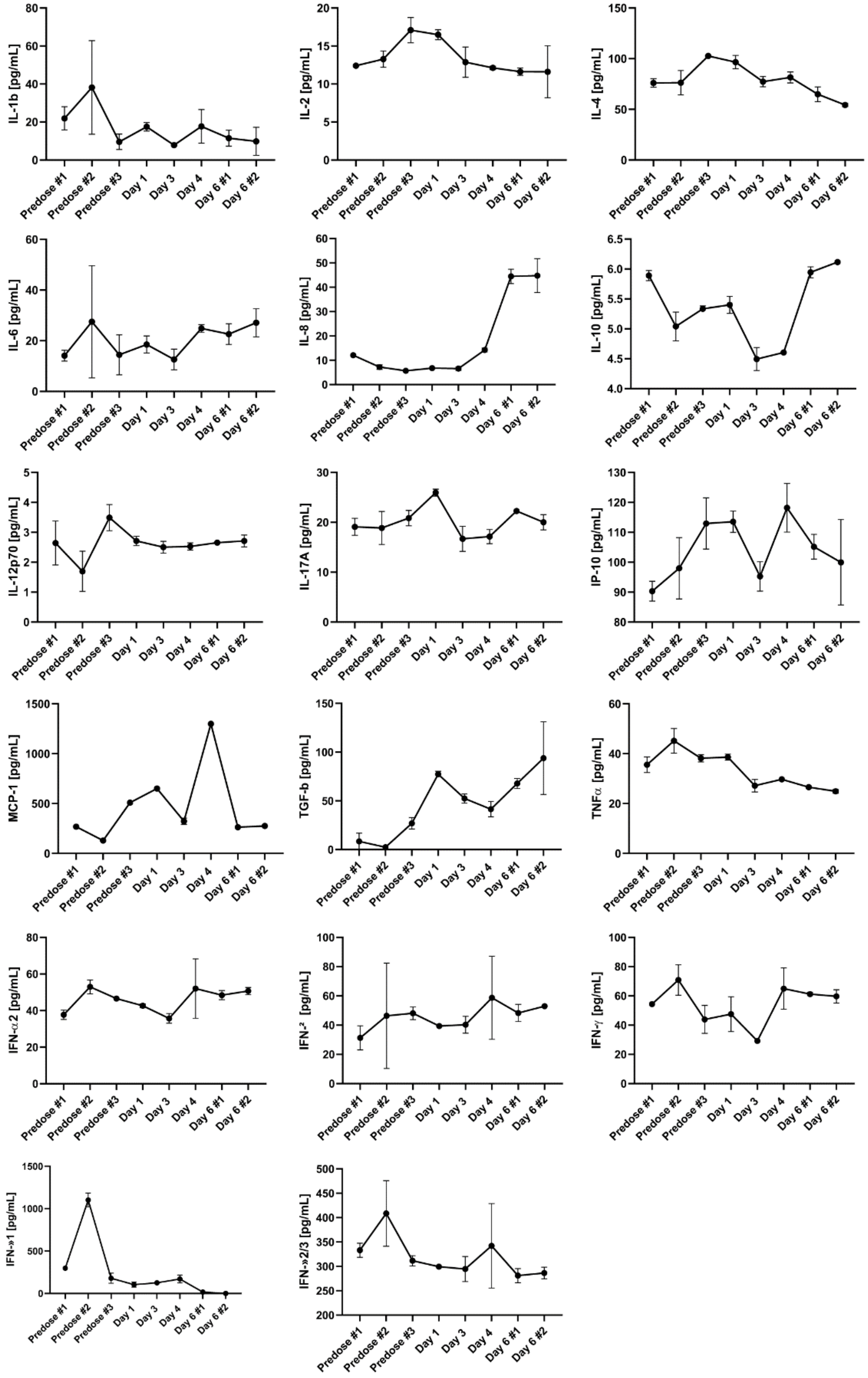

**Figure S3: Serum Cytokines over time.** Multiplex Cytokine bead assay of serum from patient before and after gene therapy. Error bars represent technical replicates.

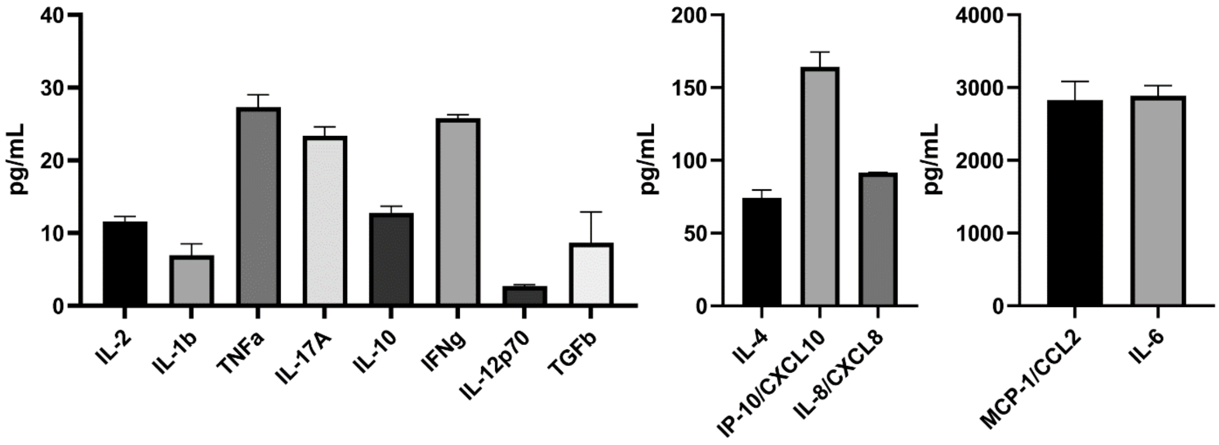

**Figure S4: Pericardial Fluid Cytokines.** Multiplex Cytokine bead assay of Pericardial fluid removed from patient day 6 post-treatment. Error bars represent technical replicates.

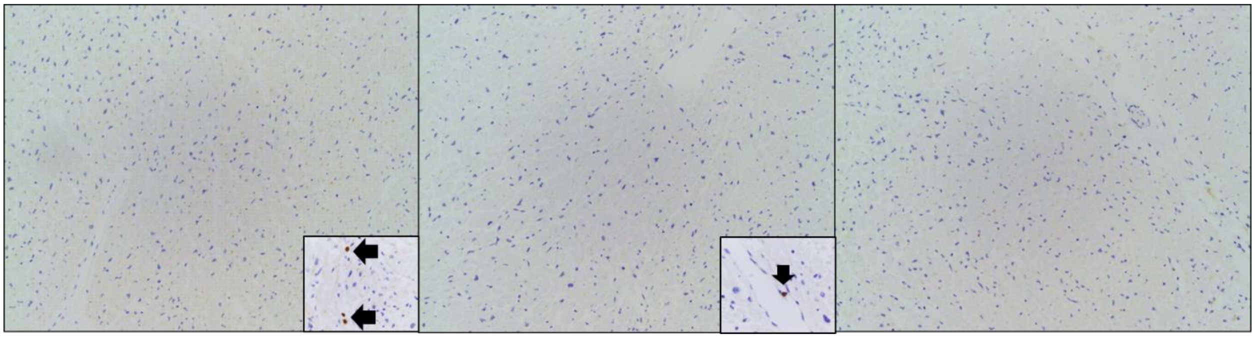

**Figure S5: Post-mortem immunohistochemistry on cardiac tissue.** There are no increased lymphocytes, either T or B cells, in the heart tissue by CD3 and CD20 immunohistochemistry, respectively (10×). Insets show internal positive control of rare intravascular T cells (CD3, 20×) and B cell (CD20, 20×), pointed out by arrows. C4d (10×) stain is also negative in the heart tissue. The findings argue against inflammatory and complement-mediated processes.

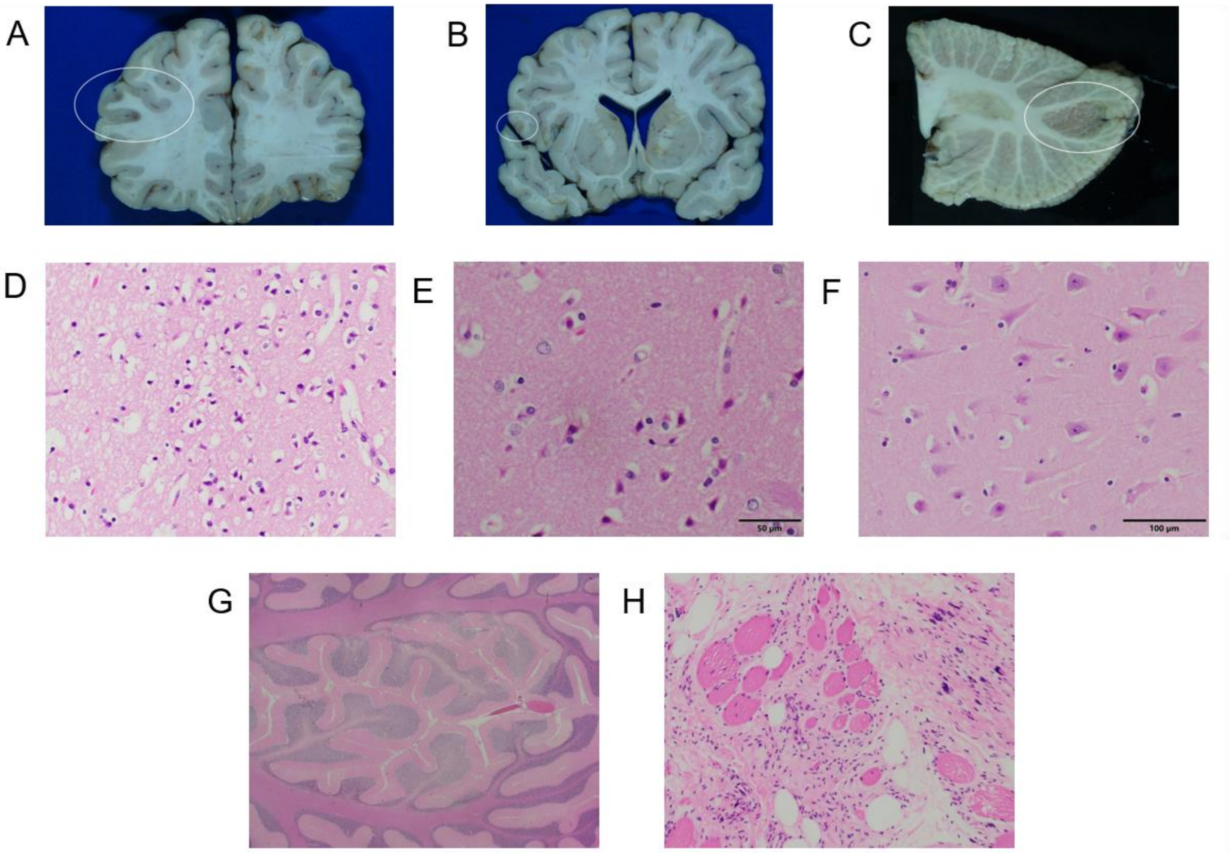

**Figure S6: Supplemental Histology Images. A)** Gross examination of a coronal section of the frontal lobe showing discolored foci (white ellipse) of infarction in the left inferior frontal gyrus. **B)** Gross examination of a coronal section of the frontal lobe showing discolored foci (white ellipse) of infarction in the left inferior frontal gyrus. **C)** Gross examination of the cerebellum showing discolored foci (white ellipse) of injury between superior and inferior cerebellar artery distributions, which is characteristic of watershed infarction. **D)** Histological analysis (40×) of the frontal cortex shows scattered neurons with dark pyknotic nuclei and markedly hypereosinophilic cytoplasm, indicating hypoxic ischemic injury days- to hours-old. Additionally, the neuropil is finely vacuolated suggesting cerebral edema. **E)** Histological analysis (60×) of the caudate nucleus shows scattered neurons with dark pyknotic nuclei and markedly hypereosinophilic cytoplasm, indicating hypoxic ischemic injury days- to hours-old. **F)** Histological analysis (40×) of hippocampal pyramidal cells shows many pale with indistinct nuclei and mild hypereosinophila, reflecting recent hypoxic ischemic injury. **G)** H&E examination (1.25×) of the focal injury seen in the cerebellum (C). Note the diffuse pallor in the central area of the cortex affecting the internal granule cell layer and, less conspicuously, the molecular layer, in contrast to the surrounding periphery. **H)** H&E examination (20×) of a paraffin-embedded section of triceps muscle showing severe fibrosis, fatty replacement, and depletion of myofibers, with the few remaining fibers being abnormally round with internalized nuclei.  A few severely atrophic fibers and "nuclear bags" are present. All features are consistent with end stage dystrophic myopathy.

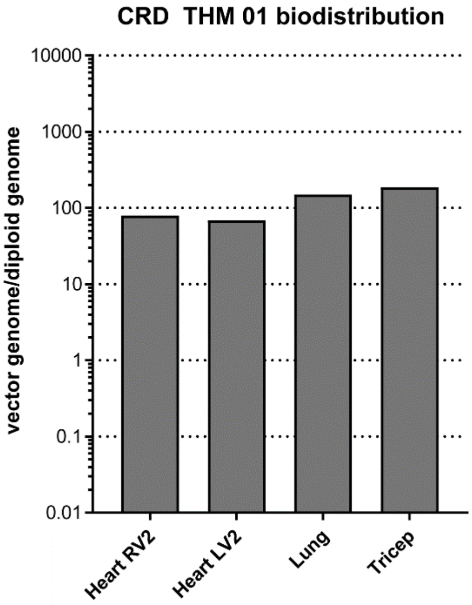

**Figure S7: ddPCR of post-mortem tissues.** ddPCR for vector genomes confirmed qPCR values for heart (RV = right ventricle, LV = left ventricle), lung and triceps muscle, but liver vector genome number was above upper limit of detection.

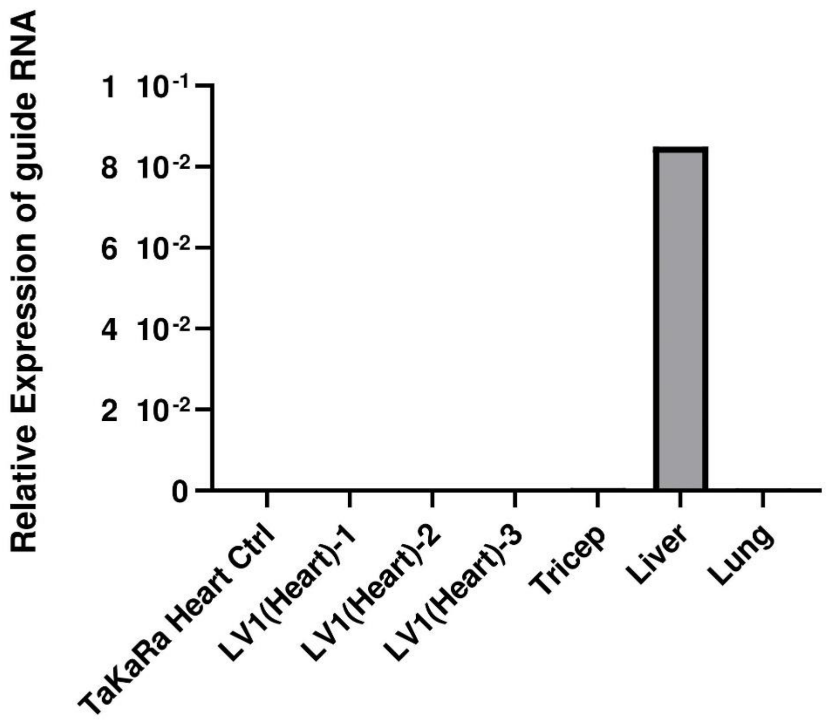

**Figure S8: Expression of guide RNA.** Expression level of the guide (C7) was measured by RT-qPCR and normalized to the housekeeping gene hHPRT1. qPCR triplicates were performed for each reaction. Values reported were quantified with ddCT. Left ventrical (LV) was sampled in 3 different places.

### SUPPLEMENTARY TABLES

#### Table S1: Certificate of analysis for GMP-grade AAV manufactured by Andelyn.

| | |
|---|---|
| **Product Name:** | CRD-TMH-001 |
| **Product Description:** | The biological product is a non-replicating, serotype AAV9 single-stranded (ss) recombinant adeno-associated virus (rAAV) designated CRD-TMH-001. The vector plasmid Construct1_singleVP64_C7 contains the transgene dSaCas9-VP64 which encodes staph aureus Cas9 protein/guide RNA sequences. Expression of the transgene cassette is under the control of the Ck8e promoter. The biologic product will be administered to enable in vivo expression of Dp427c. |
| **Lot Number:** | G0071A0122 |
| **Concentration:** | 4.07E + 13 vg/mL |
| **Date of Manufacture:** | 031122 (Vialing Date) |
| **Expiration Date:** | Pending Stability Testing |
| **Storage Conditions:** | ≤ -60 °C |
| **Yield (for Clinical Use):** | 306 Vials at 1.75 mL per vial |

| Test Description | Acceptance Criteria | Result |
|---|---|---|
| Sterility - USP <71> | No Growth or Negative | Negative |
| Bacteriostasis/Fungistasis - USP <71> | Pass | Pass |
| Endotoxin | < 2.83 EU/mL | < 0.062 EU/mL |
| Physical Titer | > 2.5E+13 vg/mL | 4.07E+13 vg/mL |
| Infectious Unit Titer | Report Result | 1.7E+09 IU/mL |
| Total Protein | Report Result | 352.711 µg/mL |
| Purity | Purity > 90% with no single impurity > 2% | Total Purity = 100%<br>Impurity Bands:<br>82.9 kD < 1%<br>76.5 kD < 1%<br>67.4 kD < 1%<br>49.9 kD < 1%<br>46.7 kD < 1%<br>43.4 kD < 1%<br>41.7 kD < 1%<br>33.9 kD < 1%<br>32.3 kD < 1%<br>31.5 kD < 1%<br>29.6 kD < 1% |
| Identity (DNA) | DNA matches the expected sequence | DNA matches the expected sequence |
| Identity (Protein) | Positive for AAV Capsid Protein | Positive for AAV Capsid Protein |
| Osmolality | 388 – 426 mOsm/kg (mOsm) | 408.7 mOsm/kg |
| pH | 8.0 ± 0.5 | pH = 7.96 |
